# Development and validation of a transcriptional signature for the assessment of fibrosis in organs

**DOI:** 10.1101/2020.03.14.20024141

**Authors:** Bin Wang, Shiju Chen, Hongyan Qian, Rongjuan Chen, Yan He, Xinwei Zhang, Jingxiu Xuan, Yuan Liu, Guixiu Shi

## Abstract

**Background:** Fibrosis in most organs has proven to be an critical factor related to high risk of morbidity and mortality, but an adequate assessment of fibrosis severity is still challenging. This study tried to evaluate fibrosis severity through a fibrosis transcriptional signature.

**Methods:** A fibrosis transcriptional signature was developed through an integrated analysis of multiple expression profiling datasets of human organs with fibrosis-related diseases. A fibrosis severity score for each sample was the calculated through gene set variation analysis (GSVA), and its role in evaluating fibrosis severity was then analyzed.

**Results:** Ten expression profiling datasets of human tissues with organ failure were integrated with robust rank aggregation method, and a fibrosis severity score consisting of 149 genes. Most of those included genes were involved in fibrogenic pathways. GSEA analysis revealed that fibrosis transcriptional signature was significantly enriched in the fibrogenic tissues. Additionally, we found that fibrosis transcriptional signature could effectively differentiate fibrosis tissues and non-fibrosis tissues.

**Conclusion:** This study developed an useful fibrosis transcriptional signature involved in fibrosis-related diseases. This fibrosis transcriptional signature is helpful in precisely evaluating the fibrosis severity in common organs at the transcriptional level.

## Introduction

Fibrosis is a hallmark of pathophysiological progress and occurs in many organs such as lung, heart and kidney[1]. Following tissue damages, the healing and regenerative processes are initiated and adaptive fibrotic remodeling are involved in the short term [2]. However, strengthened or prolonged fibrotic remodeling are pathologic and can ultimately result in progressive fibrosis and irreversible scarring[1]. Fibrosis can affect most organs such as liver, lung and skin, and fibrosis-related diseases can occur in different organs such has cardiac fibrosis, pulmonary fibrosis, liver fibrosis and kidney fibrosis [2, 3]. Recent studies have demonstrated that fibrosis and scarring are central players of the functional failure of organs [1]. Fibrosis in organ is a common character of chronic diseases, and has proven to be an critical factor determining risk of morbidity and mortality [4-6]. There are currently no specific antifibrotic agents available for most fibrosis-related diseases. Under some conditions especially the persistent chronic injury, excessive fibrosis occurs and result in the deposition of extracellular matrix, progressive scarring and organ malfunction.

Fibrosis in organs should be adequately assessed, which is valuable for the monitoring and surveillance of patients. Fibrosis in organs such as liver and lung can be assessed with either invasive biopsy or non-invasive methods [7, 8]. Nevertheless, current methods still have many limitations, which may impede their use in monitoring disease severity or progression. Non-invasive methods such as serum-based tests have less accuracy and more unreliability than those invasive biopsy [9-12]. Histopathological staging of fibrosis through invasive biopsy also has some shortcoming such as high variance and low consistency among different operators or pathologists [13-16]. These facts highlight the need for developing a accurate method to assess fibrosis severity. Therefore, studies are needed to develop some more accurate methods than histopathological examination, which is able to accurately reflect the severity of fibrosis and also help to assess the treatment response. In recent decade, the growing progress in high-throughput technology such RNA-sequencing (RNA-seq) has largely promote the studies in human diseases including fibrosis-related diseases, which is also able to provide valuable assistances in clinical diagnosis [17-19]. This study tried to evaluate fibrosis severity through a fibrosis transcriptional signature.

## Methods

### Expression profiling data

Expression profiling data from Gene Expression Omnibus (GEO) were used in this study. GEO is a public functional genomics database which provides users gene expression profiles data. Only expression profiling data from liver, lung, kidney and heart tissues of patients with fibrosis-related diseases were analyzed in our study. The following searching words were used: idiopathic pulmonary fibrosis OR lung fibrosis OR cardiac fibrosis OR renal fibrosis OR kidney fibrosis OR liver cirrhosis OR pancreatic fibrosis OR heart failure OR dilated cardiomyopathy OR advanced diabetic nephropathy OR kidney failure OR renal failure OR end stage kidney disease. To be included into this study, the dataset must contain samples from at least 5 cases and 5 controls, and the method used to perform expression profiling analysis was based on RNA-seq. Datasets analyzing samples from patients with acute organ failure were excluded.

### Data procession

Expression profiling data from GEO were downloaded and annotated if necessary. Gene expression matrix with gene symbols were then prepared for each dataset. Every dataset contained samples from organ tissues with fibrosis, and those control samples from organs without obvious fibrosis, and samples from other types of diseases were excluded. For RNA-seq data in the form of raw read count, DESeq2 was used to calculated differentially expressed genes (DEGs) [20]. For RNA-seq data in other forms such as fragment per kilobase exon per million read mapped (FPKM) or transcripts per million (TPM), limma was used for differential expression analyses [21].

### Robust rank aggregation (RRA) analysis

RRA is a good and widely used method which can integrate expression profiling data from different technological platforms in an unbiased manner [22]. To identify those genes aberrantly up-regulated in the organ tissues with fibrosis, the lists of DEGs of those included datasets were integrated with RRA method. In this study, we firstly developed a fibrosis transcriptional signature by integrating all included datasets, and those up-regulated genes with a log fold change (logFC)>1.0 and adjusted P less than 0.05 were identified as significant genes involved in the fibrosis. We also developed a another fibrosis transcriptional signature by integrating datasets with large sample size (at least 20 samples), in which those up-regulated genes with a logFC>1 and adjusted P less than 0.25 were identified as significant genes involved in the fibrosis.

### Gene set variation analysis (GSVA)

GSVA is a widely used bioinformatics method which can condense information from single sample transcriptome profiling data into an enrichment score [23]. The score of one gene set for a functional pathway or disease gene signature enriched in one sample can be calculated out and then be used to evaluate whether this gene set is aberrantly up-regulated in the sample. To evaluate the fibrosis severity quantitatively through the fibrosis transcriptional signature developed above, the enrichment score of the fibrosis gene signature in each sample was thus calculated through GSVA method. GSVA was performed using 3 datasets with large sample size, which were GSE142025, GSE124685 and GSE116250, respectively.

### Gene set enrichment analysis (GSEA)

To validate whether the fibrosis gene signature was significantly enriched in samples with fibrosis, the well-known GSEA method was further used in this study [24]. Unlike GSVA, GSEA could assess the enrichment of one gene set at the group level but not at single sample level. In the GSEA analysis, the normalized enrichment score (NES) for statistical significance was defined as more than 1.0 and false discovery rate (FDR) was defined as less than 0.25. Three datasets including GSE142025, GSE124685 and GSE116250 were analyzed using GSEA.

### Statistical analysis

To compare the difference of GSVA enrichment score between fibrosis cases and non-fibrosis controls, student’s t test was used. The diagnostic role of the fibrosis gene signature was analyzed using receiver operating characteristic (ROC) curve, and the area under the curve (AUC) was also calculated out. P value less than 0.05 was considered statistically significant.

## Results

### Characteristics of expression profiling datasets on fibrosis

A total of 10 expression profiling datasets of human tissues with organ failure met the inclusion criteria and were included in this study. The characteristics of expression profiling datasets were shown in Table 1 (Table 1). Five datasets analyzed expression profiling in the lung tissues of idiopathic pulmonary fibrosis (IPF) and non-IPF controls, 4 datasets analyzed expression profiling in the heart tissues of patients with heart failure and controls, and one dataset used kidney tissues from patients with advanced diabetic nephropathy and controls (Table 1).

**Table 1.**
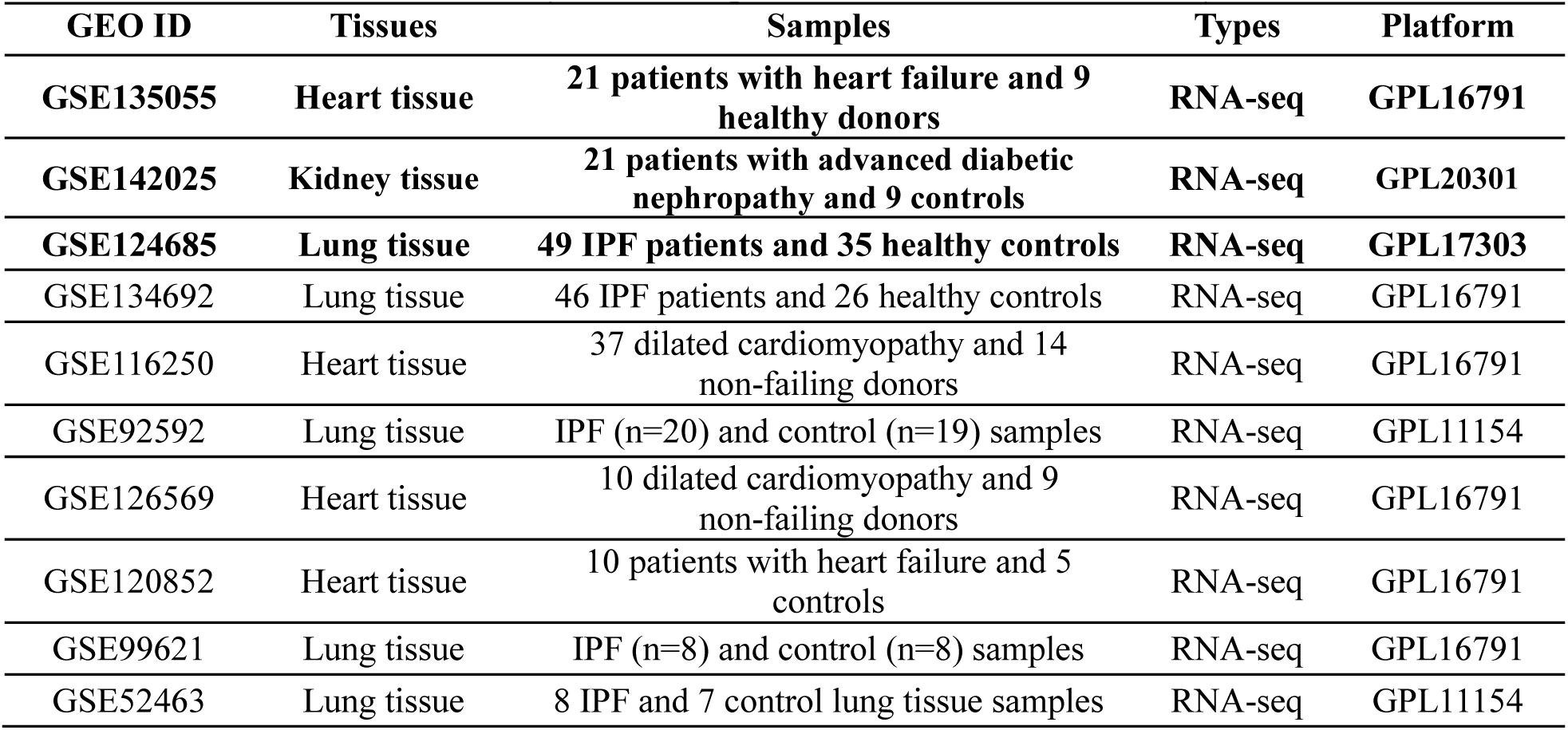
Summary of transcriptome datasets used in this study.

### Development of fibrosis transcriptional signature

We firstly developed a fibrosis transcriptional signature by integrating all 10 included datasets, and those up-regulated genes with a logFC>1.0 and adjusted P less than 0.05 were identified as significant genes involved in the fibrosis. A total of 149 genes were identified as fibrosis-related genes and were defined as a fibrosis-specific gene set named as “Fibrosis transcriptional signature” (Table 2). Those top significant genes were also shown in the Figure 1 (Figure 1). Another fibrosis transcriptional signature with less number of genes were developed by integrating datasets with 6 large sample size, which consisted of 75 genes was named as “Minor fibrosis signature” (Table 2).

**Table 2.** Fibrosis transcriptional signatures developed in this study.

| Gene set | Number | Genes |
| --- | --- | --- |
| Fibrosis transcriptional signature | 149 | COL14A1, CCDC80, ASPN, COMP, CXCL14, CPXM2, DIO2, SMOC2, MMP7, MDK, COL1A1, CFH, SSC5D, COL15A1, CTSK, THBS2, COL1A2, CTHRC1, SFRP4, GREM1, SLN, THY1, FNDC1, SULF1, KRT14, CERCAM, LUM, SFRP2, DOK5, MMP2, CPXM1, OGN, LRRC17, CPA3, FBLN2, EPHA3, CILP, CRABP2, MMP13, CCL5, FAP, FRZB, PTGFRN, COL5A1, SPP1, PROM1, SERPINF1, IGLL5, EFNB3, HDC, THBS4, BOC, LTBP2, MMP11, SLAMF7, MEOX1, PRRX2, ITGBL1, IGFL2, HBA2, LOXL1, SCARA3, CD27, IGF1, ECT2L, LY6D, HBA1, RGS4, PODN, PAMR1, ISLR, ADAMTS16, FDCSP, CLEC11A, SERPINE2, MXRA5, FCRL5, ANKRD36BP2, GPR87, IGDCC4, APLNR, ACTG2, MOXD1, CYP24A1, TSHZ2, MMP1, CP, POU2AF1, MZB1, COL3A1, DNAJC22, TUBB3, MMP10, CXCL12, VCAN, EYA2, SAMD11, BAAT, WNT10A, CD79A, TDO2, LAMP5, ALDH1A3, TNS4, DHRS9, CHST6, TFAP2A, SERPINB5, POSTN, COL16A1, BHLHE22, ADRA2A, CMA1, AEBP1, KIAA1755, CST1, MS4A1, UPK1B, IGFL1, CTSG, FMOD, UCHL1, MB, COL10A1, S100A2, CLCA2, BPIFB1, COL17A1, CD1C, UGT1A6, MFAP2, HBB, CD209, CCL22, GZMK, ADAMTS14, FCER1A, CXCL13, VTCN1, GABRP, DERL3, PDLIM4, NGFR, KRT5, CRISPLD1, TMEM59L, CST2, CCL13, FAM83A |
| Minor fibrosis signature | 75 | COMP, COL14A1, CCDC80, KRT14, COL1A1, MMP7, LY6D, COL15A1, COL5A1, PTGFRN, DOK5, SMOC2, COL1A2, CST1, LOXL1, ASPN, SFRP4, MMP13, CFH, SFRP2, THBS2, COL3A1, PROM1, IGFL2, GPR87, CTSK, DIO2, FDCSP, MMP2, MDK, TUBB3, CERCAM, GREM1, CPXM2, SERPINB4, MXRA5, SERPINF1, SPRR1B, COL17A1, CXCL6, SPP1, SLN, AEBP1, CYP24A1, ISLR, CST2, CILP, MB, CLCA2, POSTN, CCDC3, CLEC11A, LUM, MAP1A, FLNC, SERPINB3, PLA2G2A, FCRL5, SERPINB5, ANKRD36BP2, CTHRC1, LRRC17, PRRX1, CPXM1, MMP1, GABRP, FCRLA, CP, SPRR1A, CXCL14, TMEM59L, UCHL1, S100A2, ALDH1A3, NNMT |

**Figure 1.**
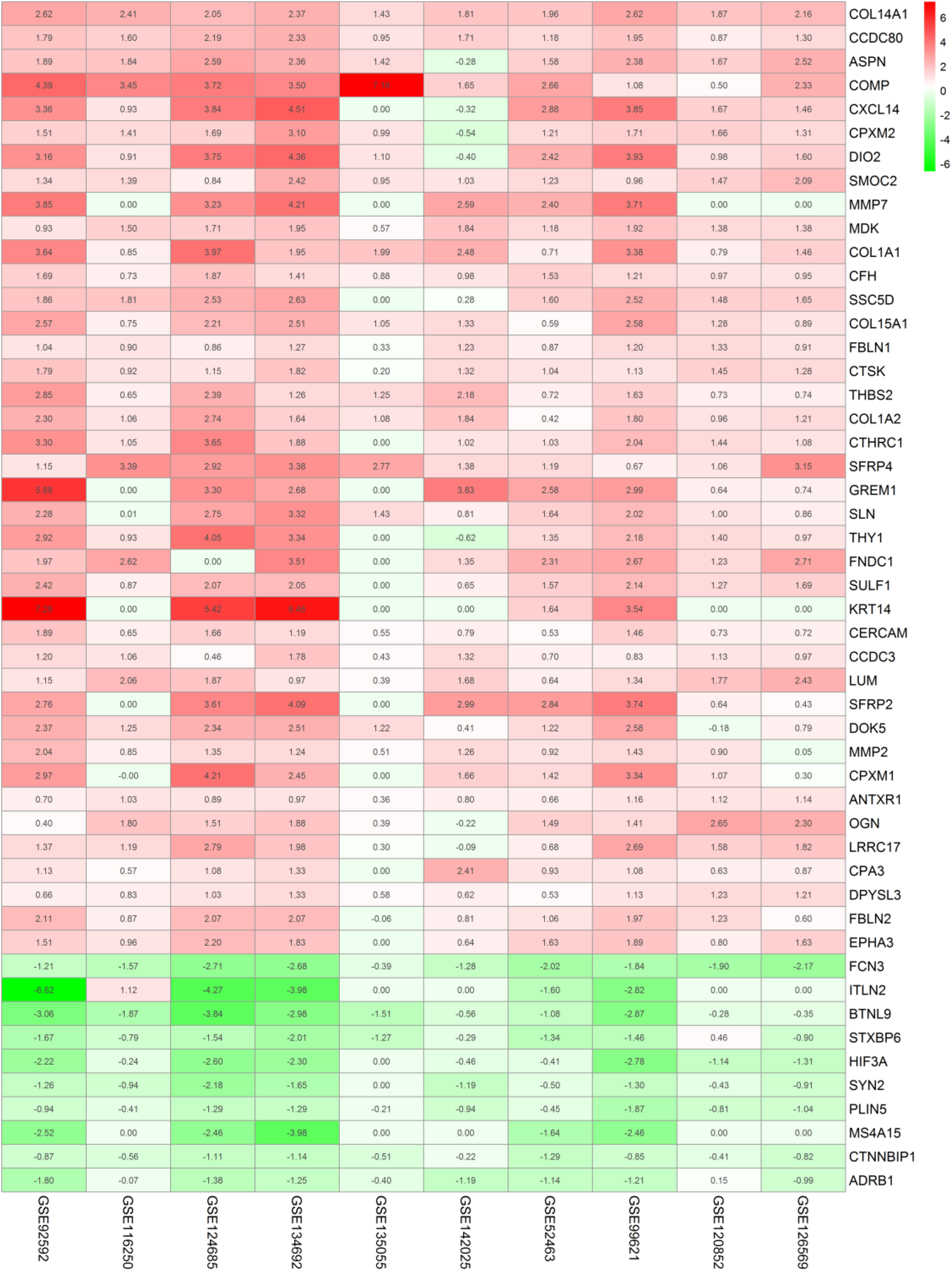
Top up-regulated genes in the fibrogenic tissues identified in RRA analysis.

### Validation of fibrosis transcriptional signature

The role of fibrosis transcriptional signature in evaluating organ fibrosis was validated through GSEA and GSVA. GSEA analysis revealed that fibrosis transcriptional signature was significantly enriched in the fibrogenic tissues from heart, kidney and lung, and it was the same with minor fibrosis signature (Figure 2 and Figure 3). Additionally, through GSVA, we found that both fibrosis transcriptional signature and the minor fibrosis signature could effectively differentiate fibrosis tissues and non-fibrosis tissues from heart, kidney and lung in the ROC analysis (Figure 4). Therefore, the outcomes above suggested this fibrosis transcriptional signature could adequately assess the severity of fibrosis in common organs at the transcriptional level.

**Figure 2.**
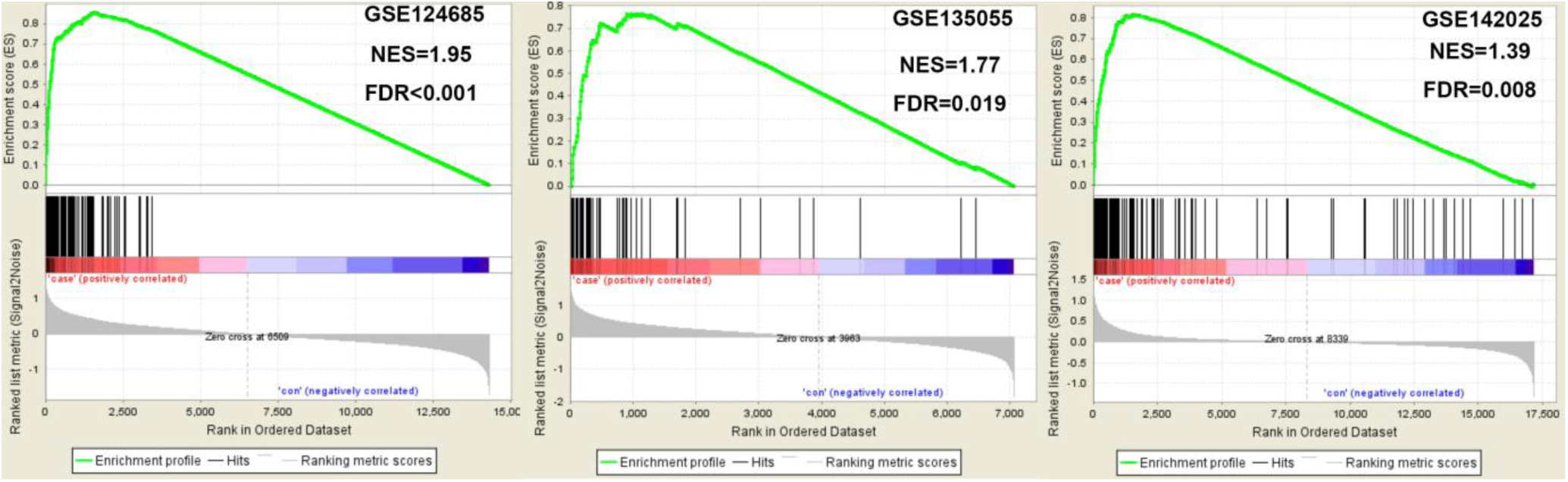
GSEA analysis revealed that fibrosis transcriptional signature was significantly enriched in the fibrogenic tissues.

**Figure 3.**
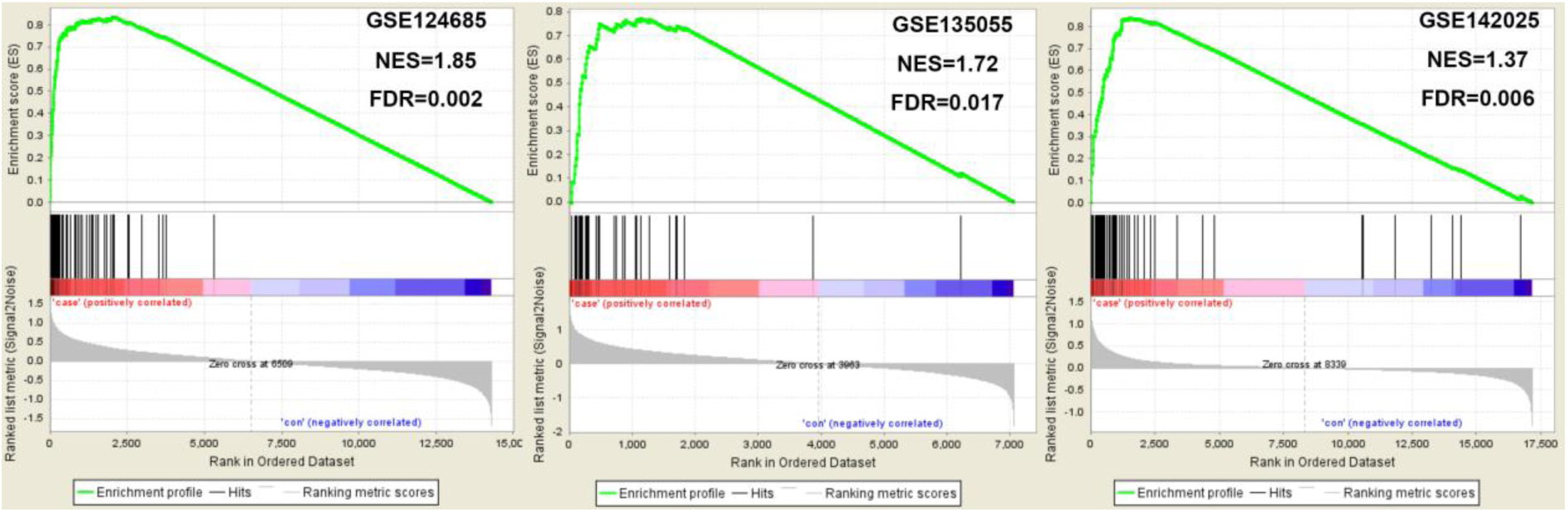
GSEA analysis revealed that minor fibrosis signature was significantly enriched in the fibrogenic tissues.

**Figure 4.**
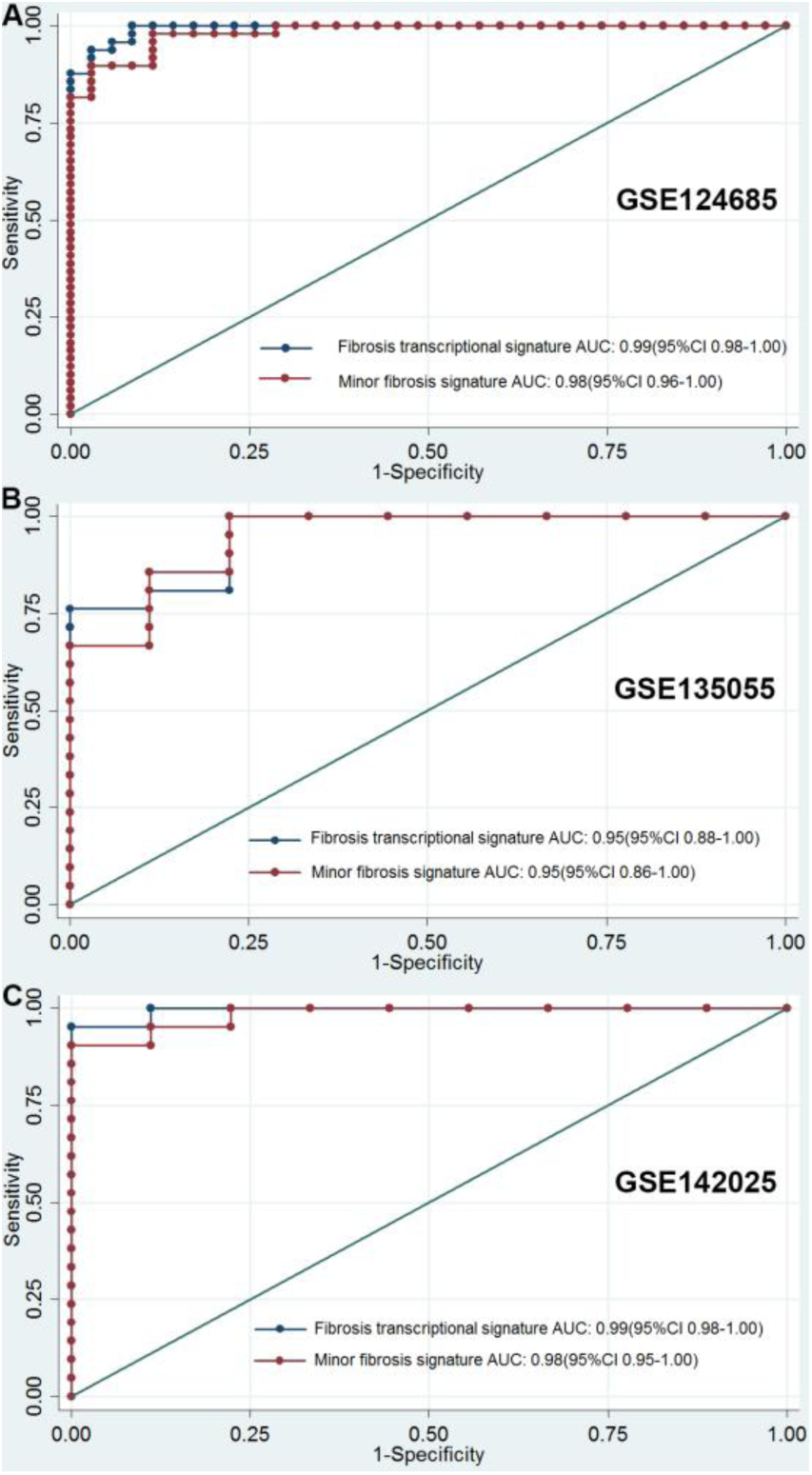
**ROC analysis suggested that both fibrosis transcriptional signature and the minor fibrosis signature could effectively differentiate fibrosis tissues and non-fibrosis tissues**

## Discussion

This study developed an useful fibrosis transcriptional signature involved in fibrosis-related diseases. To our knowledge, this is the first study aiming to develop a gene signature for the assessment of fibrosis in organs. This fibrosis transcriptional signature developed in our study is helpful in precisely evaluating the fibrosis severity in common organs such as lung, heart and kidney at the transcriptional level.

The fibrosis gene signature is a promising assessment tool which is helpful for the evaluation of disease severity at transcriptional level. Combining transcriptional gene signature and pathological examination may increases the diagnostic accuracy. Moreover, the role of fibrosis gene signature in assessing response to treatment is also important and interesting, which need to be elucidated in future studies. Further studies are needed assess whether this method can be routinely incorporated into the assessment of fibrosis in clinical practice. Apart from its role in assessing fibrosis severity in organs, another significance of the transcriptional signature is the precise estimation of fibrosis in the biomedical researches on the fibrosis-related diseases, which may become an useful biomedical tool in experimental studies.

The heterogeneity of fibrogenic diseases in different organs should not be ignored and different biological processes are involved in different fiobrosis-related diseases. Therefore, further studies aiming to develop disease or organ-specific gene signature are warranted.

In conclusion, this study developed an useful fibrosis transcriptional signature involved in fibrosis-related diseases. This fibrosis transcriptional signature is helpful in precisely evaluating the fibrosis severity in common organs at the transcriptional level.

## Data Availability

All data referred to in the manuscript were provioded.

## Acknowledgements

All authors would like to thank those researchers of those included studies.

